# Randomised controlled trial of social prescribing in schools to reduce loneliness in pupils (INACT): Trial study protocol

**DOI:** 10.64898/2026.03.11.26347953

**Authors:** Daniel Hayes, Robert J Booth, Feifei Bu, Neil Humphrey, Pamela Qualter, Lou Sticpewich, Jessica K Bone, Het Stuttard, Sophie Ellis, Sophia Maguire, Lucas Caetano Gonzalez Umpierrez, Emily Stapley, Marc S Tibber, Daisy Fancourt

**Affiliations:** Research Department of Behavioural Science and Health, Institute of Epidemiology & Health Care, Social Biobehavioural Research Group, University College London, 1-19 Torrington Place, London, WC1E 7HB; Manchester Institute of Education, School of Environment, Education and Development, The University of Manchester, Oxford Road, Manchester, M13 9PL; Evidence Based Practice Unit, University College London and the Anna Freud Centre, 4-8 Rodney St, London N1 9JH; Department of Clinical, Educational and Health Psychology, Division of Psychology and Language Sciences, 1-19 Torrington Place, London WC1E 7HB, UK

**Keywords:** Social prescribing, Wellbeing, Link worker, Loneliness, Schools, Child, Young person

## Abstract

**Background:** Social prescribing (SP) connects individuals with sources of support within their local communities and has been shown to improve loneliness. However, uptake from young people (YP) has been lower than for adults, in part due to them not accessing SP in primary care where it has been predominantly based. The INACT (INcreasing AdolesCent social and community support) programme is a novel, co-produced, SP pathway via schools for YP to access community assets and support.

**Methods:** This trial utilises a two-group (intervention vs. active control) parallel randomised design, with YP as the unit of randomisation. A minimum of 215 YP will be recruited across approximately 30 mainstream schools (primary and secondary) in England. YP reporting high levels of loneliness (7 or above on the Good Childhood index) will be randomly allocated to receive either SP or signposting. SP will consist of 6-12 sessions with a Link Worker who will work with individuals, on a one-to-one basis, to understand ‘what matters to them’ and connect them with local sources of support, whilst pupils in the signposting arm will receive a leaflet from a school staff member detailing the same sources of support. YP will be followed up at 3, 6, and 12 months after treatment allocation. Secondary outcome measures include wellbeing, emotional difficulties, service use, health related quality of life, stress, emotional regulation, as well as intervention feasibility, acceptability and appropriateness. Data about health-related quality of life and service use will be used to investigate the cost-effectiveness of the INACT programme.

**Discussion:** This trial will provide robust evidence about the effectiveness and cost-effectiveness of the INACT programme and whether it can be recommended for use in practice. The trial is due to finish 30 June 2027.

## Introduction

The transition from childhood to adolescence (with adolescence comprising the period between 10–24 years of age) is a critical developmental period, with major physical, psychological, and social changes occurring^1^. During this time, social connections, including friendships, are known to be cornerstones of healthy adolescent development^2^. Without social and community connections, negative impacts can occur^2^, including loneliness, which in turn triggers negative cycles of psychophysiological symptoms^3^. Loneliness is longitudinally associated with depressive symptoms from childhood to adolescence^4^ and can result in physical manifestations, such as poor sleep, changes to appetite, and headaches^5–7^. Young people (YP) who report loneliness are also at increased risk of cardiovascular disease and psychiatric disorders in adulthood^8,9^. It is theorised that prolonged loneliness adversely affects health via multiple mechanisms, including the dysregulation of physiological systems involved in social connections^10^.

Data from the UK show that YP report some of the highest prevalence rates of loneliness across different age groups, with 11.3% of YP aged 10-15 years old stating they always or often feel lonely^11^. This is not distributed equally, with higher levels of loneliness reported amongst younger individuals (14%), those who live in cities (19.5%) and those who receive free school meals (27.5%)^11^. In the last 30 years, data from international samples of YP suggests that levels of loneliness have been increasing, with marked rates of growth occurring from the late 1990’s onwards^12,13^. The Covid-19 pandemic further exacerbated feelings of loneliness, with 35% of YP reporting often feeling lonely during lockdowns^14^.

Recent meta-analyses of interventions that tackle loneliness in YP demonstrated small to moderate effect sizes overall^15,16^. In some instances, the authors noted that interventions targeted YP *at risk of loneliness* (e.g., due to physical health difficulties), rather than those *who reported lonelines*s^15^. Whilst across studies, authors suggested that a ‘one-size fits all’ approach is unlikely to reduce loneliness among YP, and that a tailored approach based on individual needs should be considered in future interventions. Clear evidence gaps include the lack of longer-term follow-up and insufficient consideration of socioeconomic and other risk factors^15,16^.

Social prescribing (SP) is a mechanism of care aimed to address health inequalities by linking patients with non-medical forms of existing support within their local communities^17^. It usually involves a health or social care professional referring a patient to a Link Worker (LW), who co-develops a non-clinical plan with the patient based on their values, needs and preferences and connects them with existing community organisations to improve health, wellbeing or other aspects of the patient’s life^18^. Community support can include (but is not limited to) the arts, music, access to nature, volunteering, gardening, exercise, and broader support services^17^. The future priorities of the UK’s ‘Campaign to End Loneliness’ firmly support the need for SP services, highlighting the need for ‘connector services that reach, understand, and support lonely individuals’ to ‘increase connections, improve relationships and change an individual’s thinking’^19^. These services can be delivered via psychological methods, one-to-one support, and group support. SP incorporates elements of all three^20^.

Recent reviews into the impact of SP on loneliness suggest that SP can have a positive impact^21^ and that both participants and service providers believe it to be helpful . However, those studies were limited to adult populations and often lacked control groups^22^. When control groups are utilised, results suggest that SP is beneficial^23,24^. In one UK study on SP for adults who reported loneliness, 37% of those who received SP were classified as ‘not lonely’ at 3-month follow-up, compared to 20% of those in the matched control group^24^. There was also evidence to suggest that younger age groups (those ages 18-50 years old) benefitted more than older adults^24^. Similarly, a controlled trial in Australia exploring the impact of SP on 114 adults showed that 8 weeks after starting SP, the intervention group reported decreased loneliness with a large effect size^23^. Retention of those allocated to SP was also high at 79.4%^23^ . Whilst the evidence for youth SP is less developed, a recent review found that SP is a promising way of tackling loneliness among YP, although there are methodological concerns, including small sample sizes, meaning robust conclusions could not be drawn^25^. Accordingly, there is a critical need for robust research that can provide insight into SP for YP who report loneliness.

SP is supposed to be an ‘all age’ model^26^, yet uptake from YP remains lower than for adults^27^. While YP report positive feelings towards SP and belief in its relevance and benefits^28^, the dominant model of delivery through family doctors in the UK may be inhibiting youth involvement as some research indicates they do not always feel comfortable accessing wellbeing support in this way^29^. Schools may be better placed to deliver wellbeing initiatives and targeted interventions for loneliness, as they serve as a universal point of access for YP^30^. Schools could act as a conduit to connect YP to community support, and there are already examples of individual schools who have started to act as a venue to facilitate SP in the UK^31^. The INcreasing AdolesCent social and community support (INACT) pilot aimed to test the feasibility of delivering SP for pupils who were lonely^32^. Findings indicated that the SP pathway was well received with greater reductions in loneliness for those who received SP compared to signposting. As a result, a definitive trial to test the effectiveness of INACT is now warranted. The trial described in this protocol will test INACT’s effectiveness and cost-effectiveness.

## Objectives

The primary objective is to test the hypothesis that YP reporting high rates of loneliness and who are randomly allocated to SP, will report greater reductions in loneliness at 3 months compared to a control group receiving signposting to local community activities.

Secondary objectives include testing the hypotheses that, compared to controls, the intervention group will report lower levels of loneliness at 12 month follow up, and will also report increased wellbeing at 3, 6 and 12 month follow up.

A further objective is to determine the cost-effectiveness of the INACT programme.

We will also explore, through qualitative interviews, the perceived impact of INACT, how benefits were achieved, and key considerations for its provision in schools.

## Methods and Materials Design

This manuscript describes the full study protocol for the INACT trial. Ethical approval was obtained from the UCL Research Ethics Committee on 11^th^ July 2025 (ref: 1207). Approval included detailed study procedures, intervention content, outcome measures and planned analyses. This document reflects the final prospectively defined protocol prior to outcome analysis and follows the SPIRIT checklist (see supporting information S1). Any details of protocol amendments will be submitted after approval from UCL Research Ethics Committee. INACT will adopt a two-group (intervention vs. active control) parallel randomised design, with YP as the unit of randomisation. YP allocated to the intervention arm will receive SP, while those in the control arm will receive signposting from school staff to activities in their local area. Outcomes will be assessed at baseline (T0), 3 months after being allocated to SP or signposting (T1), 6 months after being allocated to SP or signposting (T2), and 12 months after being allocated to SP or signposting (T3).

## Setting

This study will involve primary and secondary schools in England. We will recruit our sample from mainstream schools across 5 cities: London, Leeds, Sunderland, Hull and Manchester. These sites have been selected due to their (i) urbanicity, (ii) diversity in socio-economic status, and (iii) high levels of deprivation, which are known to affect community connection and loneliness^11^.

## Participants

We aim to recruit approximately 30 schools. In each primary school, up to two classes will be selected from Year 5 (where YP are aged 9 to 10 years old), whilst in secondary schools up to three classes from Years 7 and 8 (children aged 11, 12 and 13 years old) will be selected. Year 6 (children aged 10-11 years old) will be excluded from taking part because previous studies by the research team have shown that schools are reluctant to engage Year 6 students in research due to exam preparation^33,34^. Private schools will be excluded because they may have greater resources to support YP with wellbeing difficulties and do not use Unique Pupil Numbers (UPNs) linked to the National Pupil Database, which will be needed to explore factors associated with low community connection and loneliness.

In addition to YP, approximately 50 other stakeholders, namely LWs and school pastoral staff, will participate in the INACT trial. Pre-intervention, a single member of staff in each participating school will be asked to complete a school wellbeing provision survey. Typically, this will be the named mental health lead; in schools where there is no named mental health lead, it may be the member of staff with primary responsibility for personal, social and health education (PSHE) provision or pastoral support. Post-intervention, school staff who were involved in signposting and LWs who provided SP will be asked to complete a feasibility, acceptability and appropriateness survey, as well as an implementation survey detailing how long they spent with pupils signposting or engaging in SP.

Finally, a subsample of YP (n=40), and stakeholders (n=20) in the intervention arm will be asked to take part in a qualitative interview to better understand the impact of the SP and signposting pathways, as well as barriers and facilitators to engagement and implementation. Topic guides are shown in the supplementary information (S2 and S3). Purposive sampling will be employed. For YP, purposive sampling will take into account socio-demographic factors, such as age, gender, ethnicity, site, loneliness score, and the degree to which they engaged in SP (none, some, a lot – for the SP group). For LWs and school staff, purposive sampling will take into account sociodemographic factors, location, role, and types of activities prescribed (for LWs). The number of interviews has been selected to be large enough to allow for adequate “information power” and to make meaningful comparisons between SP pathway delivery and experiences in different sites^35^ but not too large to dilute an in-depth rich analysis and exploration of individual participant accounts.

## Recruitment

Our recruitment strategy will include (i) drawing on established school networks (e.g., the Schools in Mind network, containing over 30,000 schools), (ii) undertaking an extensive targeted social media campaign towards educational professionals in cities of interest; (iii) attending headteacher network meetings to promote the project; (iv) purchasing access to the Sprint education database, which contains up-to-date contact details on 570,000 teachers and senior staff at UK schools, (v) offering transparent and accessible communication regarding what school participation entails formalised in a Memorandum of Understanding (MOU), and (vi) compensation of £500 per school for administrative support.

Schools will be recruited by the methods specified above over the summer and into the Autumn term with the aim of starting SP and signposting on 3^rd^ January 2026. Schools that express an interest will be asked to fill out a MOU agreeing to participate. As interest is likely to exceed capacity, some schools will be held in reserve in case of dropout. Following recruitment of schools, YP in relevant year groups will be recruited in two stages. Firstly, schools will send letters to parents/carers of pupils in selected classes, including to those identified as persistently absent. The letter will be sent between 15^th^ September 2025 and 31^st^ October 2025 and will provide information about the study and allow parents/carers to opt their children out of participation. Secondly, for parents/carers that do not opt out their YP, the YP must assent by reading through an online information sheet and completing an online consent form before they can take part in the baseline survey. Data collection for baseline surveys and YP assent will start on 10^th^ November 2025 and be completed by 20^th^ December 2025. Thus, participant recruitment will be completed by 20^th^ December 2025. Follow up data collection will be completed by 20^th^ December 2026, and the study completion date is 30^th^ June 2027. For the qualitative interviews, separate information sheets and opt-in consent forms will be provided to parents/carers and YP who are interested in participating.

To identify eligible YP, baseline surveys will be administered to classes of pupils in relevant year groups during PSHE (or another appropriate lesson determined by the school). For pupils who are persistently absent from school, baseline surveys can be completed in their own time (e.g. at home), or with researchers or school pastoral staff at pupils’ convenience. Those who meet the inclusion criteria (a score of 7 or above on the Good Childhood Index^36^) will then be randomly allocated to receive SP (the intervention) or signposting (the active control). Random allocation will be undertaken by an independent statistician at the pupil level, stratifying for location. Given the nature of the intervention, blinding of participants or researchers (other than the INACT statistician) is not possible. As LWs will only be able to work with individuals allocated to the intervention arm, this will prevent contamination effects, and all pupils will have access to the same list of community resources. Those allocated to SP will then be contacted by a LW who will arrange a time to meet with them. For those allocated to signposting, researchers will share the names of YP with school pastoral staff who will signpost them to community resources. Any YP identified as having significant mental health issues on the Me & My Feelings questionnaire^37^ will be highlighted to pastoral staff regardless of intervention group.

Participants may discontinue SP at any time without affecting their participation in follow-up assessments. Discontinuation of intervention sessions may occur if: (1) the participant withdraws consent/assent for intervention delivery; (2) safeguarding concerns require escalation to alternative services; or (3) continued participation is deemed inappropriate by school safeguarding leads. Participants who discontinue the intervention will remain in outcome follow-up unless they withdraw from the study entirely.

YP who receive SP or signposting will then be followed up at 3, 6, and 12 months using the same measures as in the baseline survey, with additional questions on what they thought of the intervention. Therapeutic alliance with the LW will also be measured in those receiving SP. Participants will receive a £10 voucher for completing follow up measures. Those who were allocated to receive SP or signposting but chose not to receive it will still be given the option of completing follow-up questionnaires for Intention-To-Treat analysis. Feasibility, acceptability, and appropriateness measures will be administered to school staff and LWs 3 months after the intervention begins. Qualitative interviews with YP will take place at least 3 months after they have received SP and interviews with school staff and LWs will take place at least 6 months after SP has been implemented in their school. Stakeholders will be approached by the research team, taking into account socio-demographic information, loneliness scores and SP engagement to allow for purposeful sampling. All personal data will be kept on the UCL Data Safe Haven and will be stored securely under a unique numerical identifier. The UCL Data Safe Haven is a certified data transfer and storage system that meets the ISO27001 information security standard and conforms to NHS Digital’s Information Governance Toolkit.

All data collection will be completed by 20^th^ December 2026 and results are expected by September 2027.

## Sample size calculation

This trial is powered to detect a small-to-moderate standardised between-group difference in loneliness of δ = 0.32 at the primary endpoint (3 months), informed by the INACT pilot trial, which observed a standardised effect size of approximately 0.32 at 3 months^32^. Assuming a two-sided significance level of α = 0.05 and 80% power (β = 0.20), and incorporating the efficiency gain from repeated measures across three post-baseline time points (w = 3) using the pilot-derived within-person intraclass correlation coefficient (ICC, ρ = 0.13), the required post-baseline analytic sample is N = 124 participants in total (62 per arm for 1:1 allocation).

Because the intervention is delivered by LWs and outcomes may be correlated within LW caseloads, the sample size is inflated using a design effect based on an assumed LW ICC of 0.02 and an average caseload of m = 25 participants per LW. The resulting design effect is DEFF = 1 + (m − 1) × ICC = 1 + 24 × 0.02 = 1.48, giving an adjusted required analytic sample of N = 184 participants (92 per arm) at the primary endpoint. To allow for loss to follow-up by the 3-month primary endpoint, the required baseline sample is inflated to N = 215 participants in total. This corresponds to an anticipated follow-up completion rate of approximately 86% at 3 months.

Based on pilot recruitment parameters, we estimate that approximately 21% of screened pupils will meet the eligibility threshold for elevated loneliness, and that 78% of eligible pupils will provide assent/consent and complete baseline measures. Therefore, to achieve N = 215 participants at baseline, we will screen approximately 1,311 pupils in participating schools (215 / (0.21 × 0.78) ≈ 1,311).

## Intervention and control

SP is a person-centred approach to wellbeing involving the co-development of a non-clinical prescription, between an individual (in this case, YP) and LW, based on the perceived reasons for the referral and the YP’s values, needs and preferences^26^. LWs have excellent knowledge of their local areas, via community asset mapping and networking, allowing them to connect individuals with different types of support and activities.

The SP intervention used in this study will be ‘YES’: Youth Engagement in SP. YES is a tailored SP programme for YP that has been co-produced with YP to connect them with forms of support in their communities. It has been used nationally in the INACT pilot, as well as in other studies such as the Wellbeing While Waiting programme which implemented SP for YP on waiting lists for mental health treatment^38^.

The YES SP intervention consists of 6-12 sessions with a LW over an 8-week period. Sessions may take place online, via phone call, or in person. As part of the SP process, LWs will draw on psychological skills such as motivational interviewing and behavioural activation^20^ as well as employing problem solving and goal setting^39^. Following the identification of individual needs and preferences, the LW will discuss available local activities and informal sources of support with YP that best match their interests and could help them to feel more connected. Then, with the YP’s agreement, the LW will facilitate their engagement with these activities, which could be done via the LW making links with the organisation and/or accompanying the YP to the first session. All LWs will have undergone training^40^ and receive supervision. LWs will maintain structured session logs documenting session attendance, duration, mode of delivery (in person/online/telephone), and activity referrals.

YP in the control group will receive signposting to social and community activities in their local area. This will consist of school pastoral staff providing YP with a leaflet detailing information on the same local sources of support identified by the LW from asset mapping.

Participants may continue to access usual school-based pastoral care, primary care services, or community support throughout the study. No restrictions will be placed on access to external services. Use of additional services will be captured through service use measures and considered in economic analyses.

## INACT measures and outcomes

A schedule of enrolment, interventions, and assessments is outlined in Figure 1

**Figure 1.**
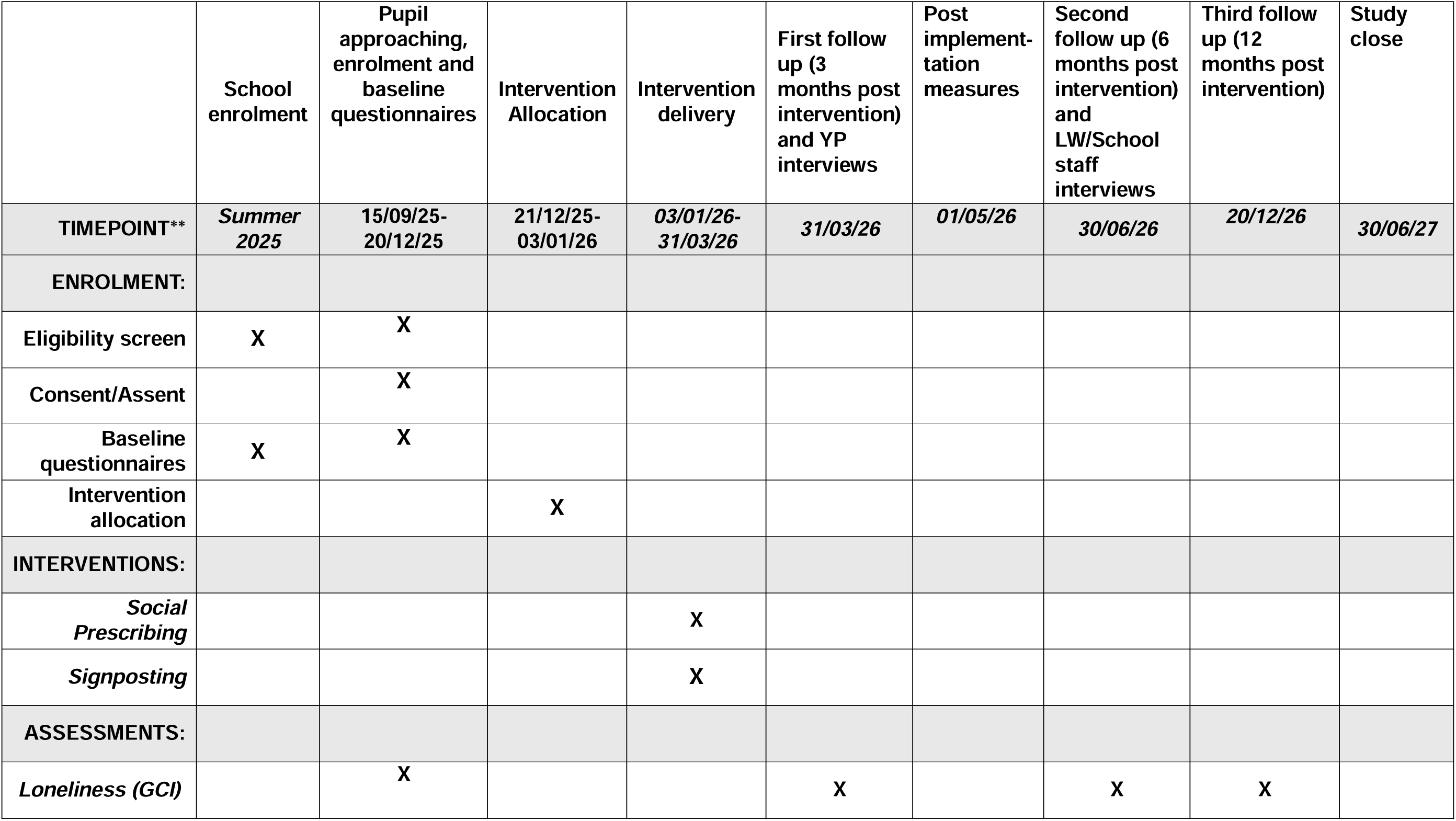
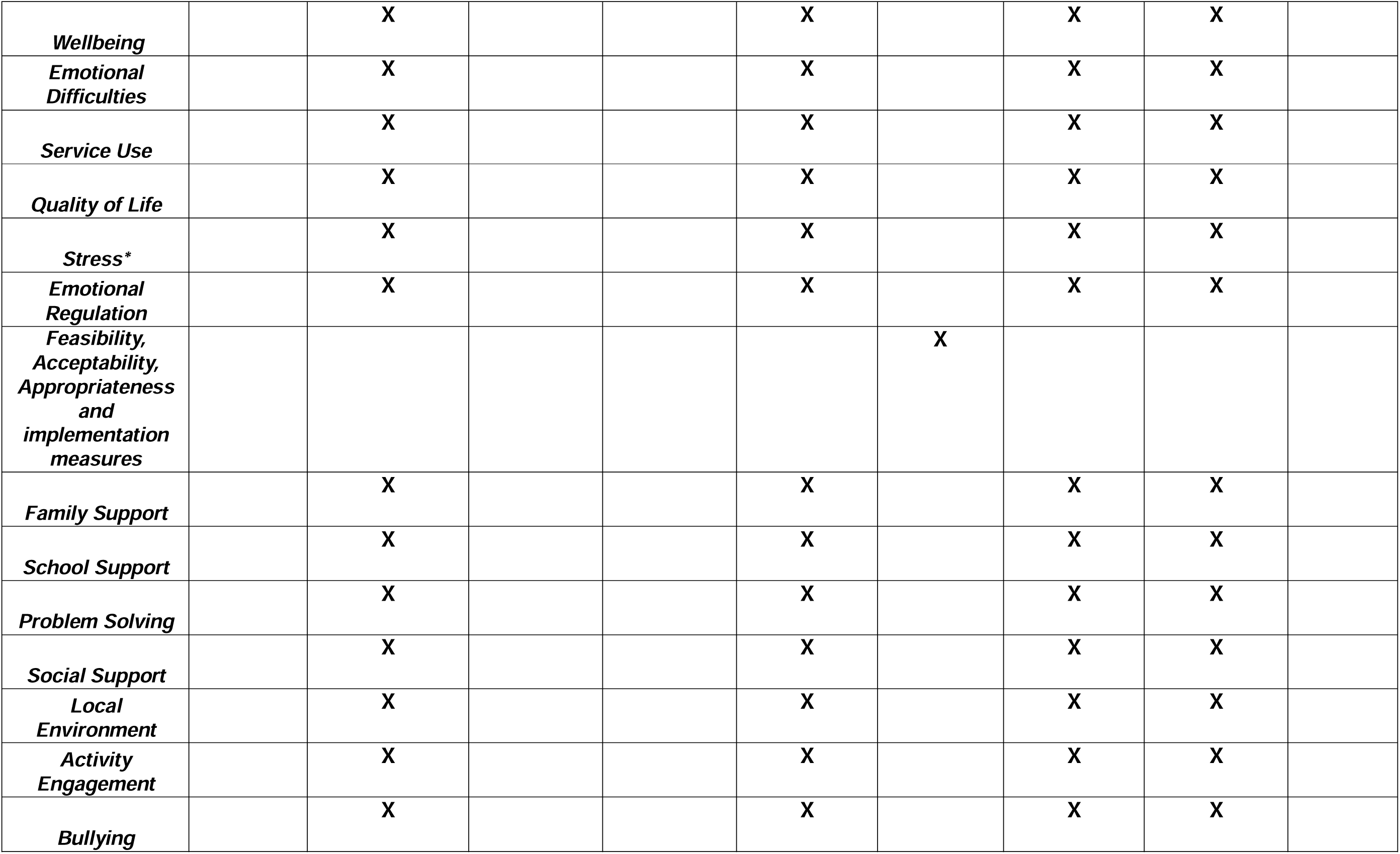
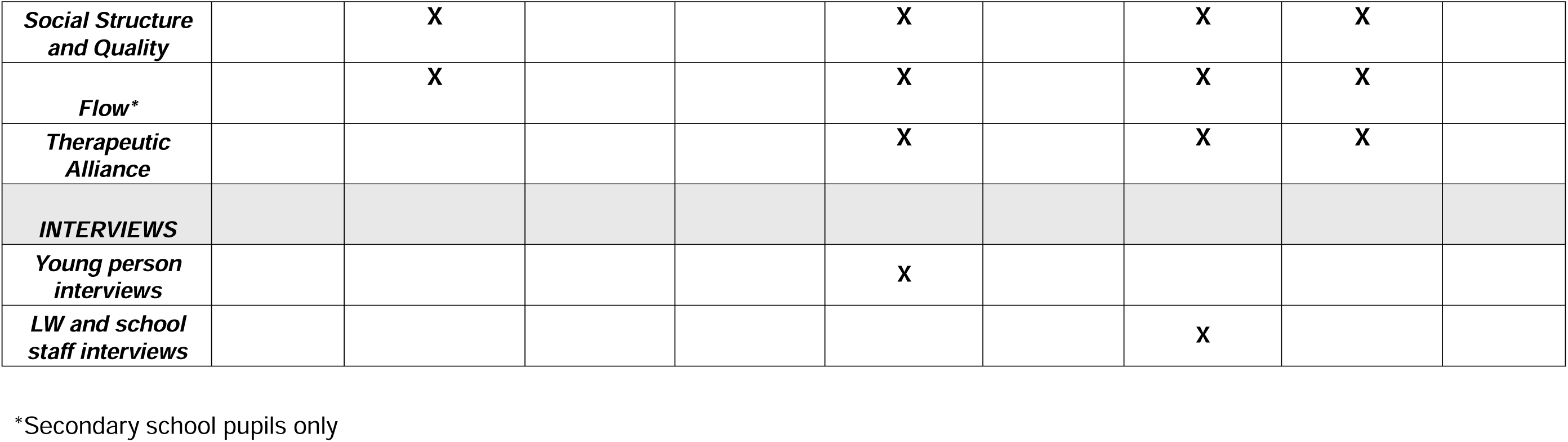
INACT Schedule of enrolment, interventions, and assessments

### Study outcome measures

The primary outcome measure for INACT is the three loneliness questions from the Good Childhood Index^36^. Each question is scored on a three-point Likert scale ranging from 1 ‘hardly ever or never’ to 3 ‘often’, resulting in total scores from 3 to 9 Higher total scores indicate higher levels of loneliness. This will be assessed at baseline, 3, 6, and 12 months, with the primary timepoint considered as 3 month follow up.

Secondary measures were chosen with YP and subject expert input. These include:

### For YP

- Wellbeing – assessed using the Wellbeing Scale from the Kidscreen-52^41^. This consists of 6 questions each on a five-point Likert scale ranging from 1 ‘not at all/never’ to 5 ‘extremely/always’. Higher total scores indicate greater well-being. This is assessed at baseline, 3, 6, and 12 months.
- Emotional difficulties – assessed using the emotional difficulties subscale from the Me and My Feelings questionnaire(Deighton et al., 2013). This consists of 10 questions each on a three-point Likert scale ranging from 0 ‘never’ to 2 ‘always’. Higher total scores indicate greater emotional difficulties. This is assessed at baseline, 3, 6 and 12 months.
- Service use – assessed using the Client Service Receipt of Inventory (CSRI; Beecham & Knapp, 2001). It contains 11 questions on a five-point Likert scale. Scoring can be looked at by individual items (i.e. score between 1-5) or by scoring all items (i.e. scores between 11-55). Higher scores indicate more contact with a service/services. This is assessed at baseline, 3, 6, and 12 months.
- Health Related Quality of Life – assessed using the Child Health Utility 9D (CHU-9D)^43^. It consists of 9 questions each on a five-point Likert Scale asking about daily life and how the individual feels that day in relation to aspects such as pain, usual activities, schoolwork/homework, tiredness, and sleep. Higher scores indicate higher quality of life. This is assessed at baseline, 3, 6, and 12 months.
- Stress - assessed using the Perceived Stress Scale^44^. This consists of 4 questions each on a five-point Likert scale ranging from 1 ‘never’ to 4 ‘very often’. Higher scores indicate higher levels of stress. This is assessed at baseline, 3, 6, and 12 months. Due to its reading age, this measure will only be used with secondary school pupils.
- Emotion regulation – assessed using the Emotion Regulation Index for Children and Adolescents^45^. This consists of 16 questions each on a five-point Likert scale ranging from 1 ‘strongly disagree’ to 5 ‘strongly agree’. Higher scores indicate higher levels of emotion regulation. This is assessed at baseline, 3, 6, and 12 months.

## Other measures

- Family support – assessed using the family support subscale from the Student Resilience Survey^46^. It consists of 4 questions each on a five-point Likert scale ranging from 1 ‘never’ to 5 ‘always’. Higher scores indicate higher family support. This is assessed at baseline, 3, 6, and 12 months.
- School support – assessed using the school support subscale from the Student Resilience Survey^46^. It consists of 4 questions each on a five-point Likert scale ranging from 1 ‘never’ to 5 ‘always’. Higher scores indicate higher school support. This is assessed at baseline, 3, 6, and 12 months.
- Problem solving – assessed using the problem solving subscale from the Student Resilience Survey^46^. It consists of 3 questions each on a five-point Likert scale ranging from 1 ‘never’ to 5 ‘always’. Higher scores indicate better problem solving. This is assessed at baseline, 3, 6, and 12 months.
- Social Support – assessed using the Child and Youth Resilience Measure^47^.

This contains 4 questions each question on a five-point Likert scale ranging from 1 ‘not at all’ to 5 ‘a lot’. Higher scores indicate greater social support. This is assessed at baseline, 3, 6, and 12 months.

- Local environment – assessed using questions from the Health Behaviour in School-aged Children (HBSC) 2022 survey^48^. It contains 4 questions each on a five-point Likert scale ranging from 1 ‘strongly disagree’ to 5 ‘strongly agree’. Higher scores indicate more positive views of one’s environment. This is assessed at baseline, 3, 6, and 12 months.
- Activity engagement - assessed using the BeeWell survey^49^. It consists of 11 questions on activity engagement each on a six-point Likert scale ranging from 1 ‘never or almost never’ to 6 ‘most days’. Each item can be individually scored, or a higher overall score indicates more frequent engagement in multiple activities. This is assessed at baseline, 3, 6, and 12 months.
- Bullying – assessed using the bullying subscale from the Kidscreen-52^41^ for primary school pupils and the bullying subscale from the BeeWell survey^49^ for secondary school pupils. For the Kidscreen-52^41^ this consists of three questions each on a five-point Likert scale, ranging from 1 ‘never’ to 5 ‘always’. For the BeeWell survey^49^ this contains 3 items each on a four-point Likert scale ranging from 0’ not’ bullied at all’ to 3 ‘a lot’. In both instances, higher scores indicate higher levels of bullying. This is assessed at baseline, 3, 6, and 12 months.
- Social structure and quality using questions adapted from the OECD Programme for International Student Assessment (PISA) 2022^50^ and Millenium Cohort Study Wave 5^51^. It consists of 5 questions asking about social support and quality. Of these, 4 questions each assess social structure and quality using a five-point Likert Scale ranging from 0 ‘I do not have any friends’ to 4 ‘every day or several times a day/a lot’. Higher scores indicate greater social structure and quality connections. The other question requires a numerical input on how many close friends the YP has. This is assessed at baseline, 3, 6, and 12 months.

Flow - assessed using the General Flow Proneness Scale^52^. It consists of 13 items each on a five-point Likert scale each ranging from 1 ‘strongly disagree’ to 5 ‘strongly agree’. Higher scores indicate a higher proneness for flow-state experiences. This is assessed at baseline, 3, 6, and 12 months. Due to its reading age, this is only being used with secondary school pupils.

- Therapeutic alliance – assessed using the Session Feedback Questionnaire(Evidence Based Practice Unit, 2012). It consists of four items each on a five-point Likert scale each ranging from 1 ‘not at all’ to 5 ‘totally’. Higher scores indicate higher therapeutic alliance. This is assessed at 3, 6, and 12 months for those in the intervention (SP) group.

### SP implementation measures

- For school staff and LWs, feasibility, acceptability and appropriateness will be assessed with the Feasibility of Implementation Measure, Acceptability of Intervention Measure and Appropriateness of Intervention Measure^54^. Each questionnaire consists of 4 questions, each on a five-point Likert scale ranging from 1 ‘Completely Disagree’ to 5 ‘Completely Agree’. Higher scores indicate greater feasibility, acceptability, and appropriateness. These will be assessed at 3 months only.
- As the above measures are not validated to be used with YP, acceptability will also be assessed by a single-item adapted NHS Friends and Family Test. This will consist of 1 question asking whether individuals would recommend SP to their friends or family, ranging from 1 ‘extremely unlikely’ to ‘extremely likely’.

## Data analysis Quantitative analysis

With appropriate permissions from the Department for Education, participant data will be linked to the National Pupil Database (NPD) using unique pupil numbers provided by schools. NPD linkage will enable the examination of socio-demographic characteristics (e.g., ethnicity, free school meal eligibility, special educational needs status) as predictors of loneliness, engagement with the intervention, and outcomes. Administrative data will be used for secondary and exploratory analyses only and will not influence eligibility, randomisation, or primary outcome assessment.

All analyses will be conducted according to the intention-to-treat principle, with participants analysed in the groups to which they were randomised, regardless of intervention uptake or completion. A detailed statistical analysis plan (SAP) will be finalised and locked prior to inspection of follow-up outcome data.

The primary analysis will estimate the between-group difference in changes in loneliness^36^ between baseline and 3-month follow-up, using linear mixed-effects or difference-in-difference modelling. The model will account for clustering of pupils within schools. Adjusted mean differences between groups will be presented with 95% confidence intervals and two-sided p-values (α = 0.05). Standardised effect sizes (Cohen’s d) will also be reported to facilitate interpretation.

Secondary outcomes (including wellbeing, emotional difficulties, stress, health-related quality of life, service use, and other psychosocial measures) will be analysed using similar methods. Repeated-measures models incorporating baseline, 3-, 6-, and 12-month assessments will be fitted to estimate trajectories over time and examine whether intervention effects are sustained beyond the primary endpoint. Group-by-time interaction terms will be included to test whether changes over time differ between study arms.

A within-trial cost-utility analysis will be conducted from a public sector perspective. Quality-adjusted life years (QALYs) will be calculated using CHU-9D utility scores. Resource use data collected through the Client Service Receipt Inventory will be combined with national unit cost estimates to calculate total costs. Incremental cost-effectiveness ratios (ICERs) will be estimated comparing SP to signposting. Uncertainty will be assessed using non-parametric bootstrapping, and results will be presented using cost-effectiveness acceptability curves.

Sensitivity analyses will include a) per-protocol analyses excluding participants who did not initiate SP, b) analyses excluding participants whose intervention extended beyond the 3-month assessment window, and, c) analyses using multiple imputation under missing-at-random assumptions to address missing outcome data. Patterns of missingness will be examined and reported.

Exploratory subgroup analyses will assess whether intervention effects vary by school phase (primary versus secondary), gender, and socioeconomic indicators (including administrative education data where available). These analyses will be considered hypothesis-generating.

Implementation outcomes (including feasibility, acceptability and appropriateness) will be summarised descriptively. Intervention uptake (defined as attendance at ≥1 session), number of sessions attended, and referral to community activities will be reported.

Exploratory analyses will examine whether intervention dosage (number of sessions attended) is associated with loneliness outcomes using complier-average causal effect modelling.

No interim efficacy analyses are planned. Safety data will be monitored descriptively in accordance with sponsor guidance. No formal adjustment for multiplicity will be applied; findings will be interpreted cautiously.

## Qualitative analysis

Semi-structured interviews will explore experiences of the SP and signposting pathways, perceived mechanisms of change, barriers and facilitators to engagement, and views on sustainability and scalability within school contexts. Interviews will be audio-recorded, transcribed verbatim, and anonymised prior to analysis.

Data will be analysed using framework analysis^55^, which allows for both deductive and inductive coding across stakeholder groups. An initial coding framework will be developed based on study aims, implementation constructs (feasibility, acceptability, appropriateness), and emerging themes identified during familiarisation with transcripts. Coding will be conducted by at least two members of the research team to enhance analytic credibility. Discrepancies will be discussed and resolved through consensus.

Codes will be organised into thematic matrices to enable comparison within and across participant groups (YP, LWs, school staff). Particular attention will be paid to identifying perceived mechanisms through which SP may influence loneliness, including relational processes, personalised activity matching, and contextual influences within schools and families.

Findings will be integrated with quantitative outcomes using a mixed-methods triangulation approach. Qualitative data will be used to interpret trial findings, contextualise effect sizes, and inform understanding of implementation variation across sites. Where appropriate, qualitative findings will inform refinement of intervention delivery and scalability considerations.

## User involvement

User involvement is central to the INACT study. The research team has already worked with North Thames Applied Research Collaborative Patient and Public Involvement and Engagement (PPIE) group, as well as the SP Youth Network’s Youth Advisory Group (YAG) to develop this protocol, both as part of the INACT pilot and full trial. There is interest from members of both groups in providing input into the INACT study, including reviewing study documents, interpretation of findings, co-authoring papers, as well as dissemination of results in creative and accessible formats (e.g., vlogs for YP). PPIE stakeholders will meet monthly at the start of the project to facilitate start up before moving to quarterly meetings.

## Ethical considerations

Ethical approval for this study has been obtained from the UCL Research Ethics Committee (project 1207). As per the INACT pilot^32^, other studies^33,34^, and the UK Department for Education’s procedures^56^, opt out consent will be sought from parents/guardians, recognising their responsibilities to their YP, whilst maintaining YP autonomy through their assent. UCL Research Ethics Committee approved the full consent procedure, including the use of opt-out parental consent. Information Sheets in age-appropriate language will be co-developed in collaboration with the YAG. Participants will have the right to withdraw at any point, without needing to give a reason. Encrypted Dictaphones or secure videocall platforms will be used for all interviews. Questionnaire data will be anonymised and qualitative data will be pseudonymised. Personal data will be stored securely on the UCL Data Safe Haven conforming to NHS Digital’s Information Governance Toolkit.

## Data Management

The ongoing conduct and progress of this study will be monitored by an independently-chaired Data Monitoring Committee (DMC) and Trial Steering Group. The DMC will include a statistician, clinician and lay member of the public all of whom have no conflict of interest with INACT. The DMC is independent of the Sponsor. The Steering Group will consist of academics, service users, clinicians and those involved in policymaking. The Sponsor may conduct periodic monitoring visits in accordance with institutional standard operating procedures. No independent external audit is planned beyond Sponsor oversight.

Access to the final trial dataset will be restricted to authorised members of the research team. De-identified data may be shared with external collaborators subject to data sharing agreements and funder approval.

## Monitoring Adverse Events

Monitoring of Adverse Events (AE), defined as a negative, emotional or behavioural occurrence, or sustained deterioration in a research participant, will be captured during INACT. This includes Serious Adverse Events (SAE) which are a threat to life: suicidal ideation, suicidal intent, hospitalisation due to psychiatric use of substances or death including suicide. Other AEs, such as violent behaviour, self-harm, physical injury, or any other event that an individual feels is important to report, will also be captured.

LWs and School Safeguarding Leads will judge whether they believe the AE is likely related to the intervention. On becoming aware of SAEs, the Principal Investigator/Trial Manager will report SAEs or AEs which are likely to be related to the intervention to University College London who is sponsoring the research, the DMC and Steering Committee. Other AEs will be collated and reported quarterly to these groups. School and research safeguarding protocols will also be followed as standard in addition to the reporting and documenting of AEs. Participants will have the right to stop participating in the intervention, and will be made aware of this by LWs and researchers when any AEs are reported. The study sponsor has insurance arrangements in place should participants be harmed as a result of the INACT trial.

## Discussion

Loneliness in adolescence is associated with poorer mental health, reduced wellbeing, and long-term social and educational disadvantage. While SP has been widely embedded within adult primary care services^57^, evidence for its effectiveness among YP remains limited, particularly within school-based pathways^25^. The INACT trial addresses this gap by evaluating the clinical and cost effectiveness of a co-produced SP model delivered through mainstream schools to pupils reporting elevated loneliness.

Building on pilot findings demonstrating feasibility, acceptability and preliminary evidence of impact at 3 months^32^, this trial is powered to detect a small-to-moderate effect on loneliness at the primary endpoint. The study extends previous work^15,16^ by incorporating longer-term follow-up (6 and 12 months), a formal economic evaluation, and detailed implementation and process measures. This will allow assessment not only of whether SP reduces loneliness, but also whether effects are sustained, cost-effective, and deliverable at scale within school systems.

The intervention model emphasises relational continuity, personalised goal setting, and supported engagement with community assets. The inclusion of mixed-methods evaluation will enable examination of potential mechanisms of change, including relational processes, behavioural activation, and improved access to social opportunities. Qualitative findings will contextualise quantitative outcomes and help identify factors influencing variation in engagement and impact across sites.

The trial also incorporates administrative education data linkage, enabling examination of socioeconomic and contextual predictors of loneliness and intervention engagement. This will support assessment of equity in reach and outcomes, addressing concerns that social interventions may inadvertently widen inequalities if uptake differs across demographic groups.

If effective, INACT will provide robust evidence to inform commissioning decisions regarding school-based SP pathways. Findings will contribute to the developing evidence base on non-clinical interventions for youth loneliness and may inform integration of social approaches within broader mental health prevention strategies. Conversely, if no effect is observed, the study will provide important evidence to refine or reconsider the use of SP models in this population.

In summary, the INACT full trial represents a rigorous evaluation of a school-based SP pathway for YP experiencing loneliness. By combining effectiveness, economic, implementation, and equity analyses, the study aims to generate actionable evidence to guide future policy and practice. Findings will be communicated via the UCL Social Biobehavioural group website (https://sbbresearch.org/) as well as via conferences, academic papers and via schools and service user groups.

## Sponsor

University College London is the study sponsor. The sponsor can be contacted at: Office of the Vice-Provost (Research), University College London, 2 Taviton St, London WC1E 6BT. The study Sponsor reserves the right to audit INACT at any point during, or after the trial has ended.

## Author contributions

*Conceptualization*: Daniel Hayes, Daisy Fancourt. *Funding acquisition*: Daniel Hayes, Daisy Fancourt, Alexandra Burton, Feifei Bu, Neil Humphrey, Pamela Qualter. *Methodology*: Daniel Hayes, Alexandra Burton, Feifei Bu, Neil Humphrey, Pamela Qualter, Emeline Han, Lou Sticpewich, Joely Wright, Jessica K. Bone, Sophia Maguire, Lucas Caetano Gonzalez Umpierrez, Emily Stapley, Daisy Fancourt. *Project administration*: Daniel Hayes, Robert J Booth, Emeline Han, Lou Sticpewich, Joely Wright, Sophia Maguire, Lucas Caetano Gonzalez Umpierrez, Marc S. Tibber. *Supervision*: Robert J Booth, Feifei Bu, Jessica K. Bone, Emily Stapley, Marc S. Tibber, Daisy Fancourt. *Writing – original draft*: Daniel Hayes. *Writing – review & editing*: All authors

## Supporting information

- SPIRIT checklist (S1)
- Topic guide for YP (S2)
- Topic guide for staff (S3)

## Data Availability

No datasets were generated or analysed during the development of this protocol. An anonymised quantitative dataset generated and/or analysed during the current study will be available in 2027 once the study has finished. A decision regarding storage location is yet to be finalised, please contact the Principal Investigator (DH) for further information.

