## Supplementary material for "Randomised controlled trial of social prescribing in schools to reduce loneliness in pupils (INACT): Trial study protocol": S2

S2 INACT topic guide for young people

**Topic guide** **for young people**

**INcreasing AdolesCent social and community supporT (INACT)**

**A. Opening**

Thank you for agreeing to take part in this interview. I would like to ask you some questions about your experience of meeting with your school staff (for signposting) or link worker/social prescriber (for social prescribing) and any activities that you might have attended after this meeting. The interview should last less than an hour. If it is okay with you, I would like to record this interview. Do you have any questions before we begin?

**B. Social and community support before intervention**

1. Thinking about before you started the project, what types of social and community support did you have/use? *(PROMPT: activities, groups, people you would talk to)*

**C. Research**

1. Can you please tell us how you found completing the research and the survey design? (Prompts: what did you think of the survey questions)

**D. Acceptability, feasibility and suitability of intervention**

**Meetings with the school staff/social prescriber**

1. Could you tell me about your experience of meeting with the school staff/social prescriber? *(PROMPT: What did you like/enjoy about it? What did you not like/enjoy about it? Was there anything you thought could have been done better?**)*
2. Did you find it easy or difficult to attend the meeting(s) with the school staff/social prescriber? (*PROMPT: Were you able to attend the meeting(s)? What made it easy or difficult to attend the meetings?* *Were the meetings online or in-person and how did you feel about that?)*
3. Did meeting with the school staff/social prescriber help how you have been feeling at all? (*PROMPT: If yes, why do you think it helped? If no, why not? What did you find most/least helpful?)*

**Social and community support after intervention**

1. Could you tell me about any activities or groups you joined after meeting the school staff/social prescriber? *(PROMPT: types of activities e.g., football, art lessons)*
2. Did you enjoy <activity>? (*PROMPT: Why did you enjoy/not enjoy it? Were there any specific parts of <activity> that you enjoyed/did not enjoy? Was there anything that could have been done differently to make <activity> more enjoyable for you?)*
3. Did you find it easy or difficult to attend <activity>? (*PROMPT: Were you able to go to all the activity sessions? What made it easy or difficult to attend <activity> e.g., timing and location of sessions? Was there anything that could have been done differently to make it easier for you to attend the sessions?*
4. Did you find <activity> helped how you have been feeling at all? (*PROMPT: Why do you think it helped/did not help? Were there any specific parts of <activity> that you found helpful/not helpful?) Was there anything that could have been done different to make <activity> more helpful for you?*

**E. Intention to adopt and sustained use/engagement**

1. Since starting this project, what types of social and community support do you have/use? *(PROMPT: activities, groups, people you would talk to – are these different to the types of support you had/used before the project?)*
2. Is there anything you have taken from <activity> that you use, or plan to use in your everyday life? *(PROMPTS: If yes, what is it and when/how might you use it? If no, why not? What do you think might be the challenges (if any) in using <activity> in your everyday life?)*

**F. Closing** 

We have come to the end of the interview. Is there anything else that we have not covered that you would like to add?

Thank you very much for taking the time to speak with me today.
