## Supplementary material for "Randomised controlled trial of social prescribing in schools to reduce loneliness in pupils (INACT): Trial study protocol": S3

S3 INACT topic guide for staff

**Topic guide for school staff/social prescribers**

**INcreasing AdolesCent social and community supporT (INACT)**

**A. Opening**

Thank you for your time today. I would like to ask you some questions about your experiences of [signposting (for school staff) or connecting (for social prescribers)] young people to social and community support as part of our research study. The interview should last less than an hour. If it is okay with you, I would like to audio record this interview. Do you have any questions before we begin?

**B. Professional background**

1. Can you tell me a bit about your background and experience? *(Prompts: of working as a school staff/social prescriber, working with young people)*

**C. Research**

1. Can you please tell us how you found completing the research and the survey design? (Prompts: what did you think of the survey questions)

**D. Delivery**

1. Could you tell me about your experiences of delivering the signposting/social prescribing sessions to young people as part of this study? [*Prompt: describe a typical session, what worked well, what were the challenges, barriers/facilitators to delivering content, were you able to engage with the young people*]
2. Can you talk about a session that you felt went well? *[Prompt: what went well, why, what could have been done differently?]*
3. Can you talk about a session that was more challenging for you? *[Prompt: why did it not go well, what could have been done differently, what might have helped you?]*
4. Is there anything you might want to change about the delivery of the sessions to make them work better? *[prompts: format, length, content, delivery method, work better for you, work better for young people]*

**E. Perceived impact and unintended consequences**

1. What impact, if any, do you think your sessions had on the young people who took part? What was it about the sessions that made that impact? *[Prompt: What are your views about using signposting/social prescribing to help young people who report low community connections or feel lonely]*

**F. Intention to adopt and sustained use**

1. How willing would you be to continue/adopt signposting/social prescribing for young people who report low community connections or who feel lonely in the future? (*Prompts: Why, why not, what kinds of changes would you need to make signposting/social prescribing for young people work effectively in your setting? Are there any elements that should be kept the same?)*
2. Going forward, what might be the potential barriers/facilitators, if any, to the continued use or availability of signposting/social prescribing for young people who report low community connections or who feel lonely?

**G. Closing**

We have come to the end of the interview/focus group. Is there anything else that we have not covered that you would like to add?

Thank you very much for taking the time to speak with me today.
